# Maternal iron depletion trajectories during pregnancy and postpartum and their relationship with infant birthweight: A longitudinal cohort analysis

**DOI:** 10.64898/2026.03.30.26349718

**Authors:** Parisa Kabir, Fanny Sandalinas, Rhonda Bell, Stephane Bourque

## Abstract

**Background:** Maternal iron requirements increase substantially during pregnancy, and ferritin concentrations typically decline as gestation progresses. However, the physiologic significance of this decline remains uncertain, and whether reductions in maternal iron stores relate to birth outcomes is unclear.

**Objectives:** To examine associations between maternal ferritin trajectories during pregnancy and postpartum and infant anthropometric outcomes.

**Methods:** We conducted a secondary longitudinal analysis of 1,496 mother–infant pairs from the Alberta Pregnancy Outcomes and Nutrition cohort. Serum ferritin was measured longitudinally in the second and third trimesters and at three months postpartum, with limited first-trimester data available. Values below 15 µg/L indicated iron deficiency. Multivariable linear regression assessed associations between inflammation-adjusted third-trimester serum ferritin and infant birthweight and length. Change in serum ferritin between the second and third trimesters (Δ ferritin) was examined as a marker of late-gestation iron mobilization. Postpartum serum ferritin was modelled using restricted cubic splines to account for nonlinear associations with birth weight and length.

**Results:** Ferritin concentrations declined progressively across pregnancy, with 61% of women classified as iron deficient in the third trimester. Lower inflammation-adjusted third-trimester ferritin was associated with higher birthweight, corresponding to approximately 84⍰g higher birthweight per 2.7-fold decrease in ferritin (p < 0.001). Women experiencing the largest decline in ferritin between the second and third trimester delivered infants approximately 155 g heavier than those with minimal change (p = 0.001). Higher birthweight was associated with greater odds of postpartum iron deficiency (OR per 1 kg = 1.83; 95% CI: 1.12–2.99).

**Conclusions:** In this healthy cohort, maternal iron depletion in late pregnancy was associated with higher birthweight, consistent with preferential fetal iron transfer. Women delivering larger infants exhibited higher odds of iron deficiency, suggesting sustained maternal iron depletion following greater fetal iron accretion.

## Introduction

Iron deficiency (ID) remains one of the most widespread micronutrient deficiencies, affecting an estimated two billion people worldwide (1,2). Its distribution is unequal, with women experiencing higher levels of ID than men globally (3). Although ID is most prevalent in low- and middle-income countries, it also persists in high-income countries, where recent analyses show no improvement in prevalence between 1990 and 2019 (1,4).

During pregnancy, iron requirements increase substantially to support maternal plasma volume expansion, erythropoiesis, and foetal and placental growth. ID, ID anaemia (IDA), or anaemia without concurrent ID during pregnancy are associated with multiple adverse infant outcomes, including preterm birth, low birth weight (9,10), small-for-gestational-age (SGA) (11), reduced head circumference (12), shorter birth length and stunting (13), and impaired immune function (10).

However, important gaps remain in understanding when iron insufficiency is most consequential and how dynamic changes in maternal iron stores relate to foetal growth. Longitudinal evidence from a large Irish cohort shows that ID increases sharply as pregnancy progresses, with more than 80% of otherwise healthy women classified as deficient by the third trimester (14). It remains unclear whether this pattern reflects expected physiological adaptations of pregnancy, such as hemodilution secondary to blood volume expansion, or true maternal depletion of iron stores by late gestation. Ferritin concentrations decline partly as a result of plasma volume expansion, and lower ferritin values during pregnancy are not necessarily indicative of anaemia or of iron deficiency (15). However, a recent analysis of NHANES data that incorporated additional physiological indicators of iron status suggested that the ferritin threshold used to define iron deficiency in the third trimester may need to be higher than currently recommended (20 µg/L instead of 15 µg/L). This adjustment would further increase the estimated prevalence of iron deficiency (16). Importantly, because most studies examining iron deficiency during pregnancy do not include birth outcomes or postpartum ferritin assessments, the clinical implications of declining ferritin concentrations across pregnancy remain uncertain (10).

This analysis build on previous work from the Alberta Pregnancy Outcomes and Nutrition (APrON) cohort, which found that higher maternal serum ferritin concentrations were associated with lower birth weight and head circumference in a sex-specific manner (17). In the present study, we extend that work by examining (1) maternal iron status across all three trimesters of pregnancy and at three months postpartum, (2) changes in ferritin concentrations between mid- and late pregnancy, and (3) associations with infant birthweight and birth length. By integrating longitudinal ferritin measurements with postpartum assessments, we aim to clarify the timing, pattern, and physiological significance of ID during pregnancy, and to determine whether postpartum ferritin reflects persistent iron alterations in maternal iron stores.

## Methods

### Study design and recruitment

This is a secondary data analysis of the longitudinal APrON cohort study, based in Calgary, Alberta, Canada. The APrON cohort was established in 2009 and recruits pregnant women at varying gestational ages, with follow-up at each trimester of pregnancy and up to three years postpartum. Methods and rationale of the APrON study have been detailed elsewhere (18). The APrON cohort has approximately 2200 mother-child pairs. The primary aims of the APrON study are [1] to explore the relationships between maternal nutrition status throughout pregnancy and postpartum and [2] to explore the effects on maternal mood on birth outcomes and infant development.

Inclusion for the current analysis required availability of infant birth anthropometry and at least one maternal serum ferritin measure during pregnancy. Analyses of change in ferritin between mid- and late pregnancy were restricted to women with serum ferritin measured in second trimester (T2; 14–26 weeks) and third trimester (T3; 27–42 weeks). Because first-trimester (T1; 0-13 weeks) ferritin was sparsely measured, T1 was used descriptively only.

### Ethics

All participants provided informed consent prior to being included in the study. The project was approved by the University of Calgary Health Research Ethics Board and the University of Alberta Health Research Ethics Biomedical Panel. The present secondary analysis examining maternal iron status during pregnancy and postpartum in relation to birth outcomes was approved by the London School of Hygiene & Tropical Medicine (LSHTM) Ethics Committee (30572/RR/34755, April 2024).

### Outcomes

The primary outcome variables were infant anthropometrics of weight and length, measured at birth and three months post-partum. Weight was measured in grams (g) at birth and kilograms (kg) at postpartum.

### Exposure: iron status

The primary predictor variable was maternal serum ferritin at late gestation (T3). ID was defined using serum ferritin according to World Health Organization thresholds for pregnancy (<15 µg/L).

To visualize iron fluctuations during pregnancy in greater detail, we further categorized women into ferritin groups: <15 µg/L, 15–29.9 µg/L, 30–44.9 µg/L, and ≥45 µg/L. These categories were used solely to illustrate longitudinal ferritin trajectories and were not used to define clinical iron deficiency in regression analyses.

### Inflammation and ferritin adjustment

C-reactive protein (CRP) was only available in T3, and therefore, only these ferritin data are adjusted for the effect of inflammation with the BRINDA regression method (19).

### Change in ferritin from T2 to T3

To quantify late-gestation iron dynamics, we computed Δlog-ferritin (T2–T3) for women with paired measures. This metric is interpretable as a fold-change: a 1-unit decrease in Δlog corresponds to a 2.7-fold decline in ferritin from T2 to T3. Positive values indicate a fall in ferritin across late pregnancy.

### Covariates

Potential confounders were selected a priori based on biological plausibility and included: maternal age (years), pre-pregnancy BMI (kg/m^2^), number of pregnancies (count), ethnicity (white vs non-white), education (≤high-school/technical vs university/post-graduate), household income (<$70,000 vs ≥$70,000 CAD), site (Calgary vs Edmonton), gestational age (weeks), smoking, drinking and use of substances before and during pregnancy (use vs non-use). To avoid multicollinearity, we retained ethnicity and education (and excluded income as collinear) in the final models; model diagnostics confirmed VIF <3 for all retained predictors.

### Statistical analysis

Initial descriptive statistics were reported as mean, median, or percentage. Continuous variables were examined for normality, and a skewness value between ±2 was considered normally distributed (20). Variables that did not meet these requirements were logarithmically transformed.

To describe the associations between maternal ferritin status and maternal covariates, Pearson’s Chi-square test was used if the covariate was categorical and the one-way Analysis of Variance (ANOVA) test was used if the covariate was continuous.

Multiple linear regression was used to test the association between maternal iron status and infant anthropometric outcomes (IAOs). Unadjusted and adjusted models are reported, with the final model adjusting for potential confounders identified in bivariate analysis. For both bivariate and multivariate analysis, a p-value of <0.05 was considered statistically significant.

### Ferritin dynamics models

Among women with paired T2 and T3 ferritin, we modelled birth weight and length on Δlog-ferritin (T2– T3). As we examine a biological mechanism rather than an epidemiological association, we started by not including any of the identified confounders in this model, and present unadjusted and adjusted models. As no established clinical threshold exists for defining clinically meaningful reductions in ferritin during pregnancy, we categorised Δlog ferritin into three groups representing (i) positive changes or no changes in iron stores (Δlog-ferritin (T2–T3)≤ 0), (ii) a modest reduction in iron stores (Δlog-ferritin (T2– T3)>0 and <1.5), and (iii) a large reduction in iron stores (Δlog-ferritin (T2–T3) ≥1.5). The threshold of 1.5 log-units provided a clear and interpretable separation between modest and large declines, and corresponded to the upper tail of the distribution

### Postpartum ferritin models

Initial exploratory diagnostics using restricted cubic splines (df⍰=13) indicated that the association between postpartum ferritin and birth anthropometric outcomes was non-linear. Therefore, in addition to spline modelling and to facilitate clinical interpretation, we analysed postpartum ferritin as a categorical variable (iron-deficient vs non-deficient). To examine whether infant size was statistically associated with postpartum maternal iron status, we modelled postpartum ID as the outcome in a logistic regression adjusted for maternal age, gestational age, pre-pregnancy BMI, education, ethnicity, and study site. Because postpartum ferritin is influenced by postpartum inflammation, and CRP data were not available post-partum, these analyses were treated as exploratory and interpreted with caution.

## Software

Analyses were conducted in R 4.5.2. Key packages included splines (restricted cubic splines), stats (lm), and car (VIF).

## Results

### Description of the population

A total of 1,496 maternal participant-infant pairs were included in the analysis (Figure 1). Participants had a mean age of 31.7 years and a mean pre-pregnancy BMI within the normal range. Most participants were white, well educated, higher income, and resided primarily in Calgary. Median ferritin declined progressively across pregnancy, with the greatest reduction observed between T2 and T3 (Table 2). At T3, 61% of women were classified as ID (serum ferritin <15 µg/L), 68% after adjustment for CRP (Figure 2). Anaemia prevalence was low across all trimesters and post-partum (less than 5%). At three months postpartum, ferritin concentrations increased compared to late pregnancy and approached levels similar to those observed in the T1 (Table 2). CRP concentrations were low at all time points, indicating minimal inflammation.

**Table 1.**
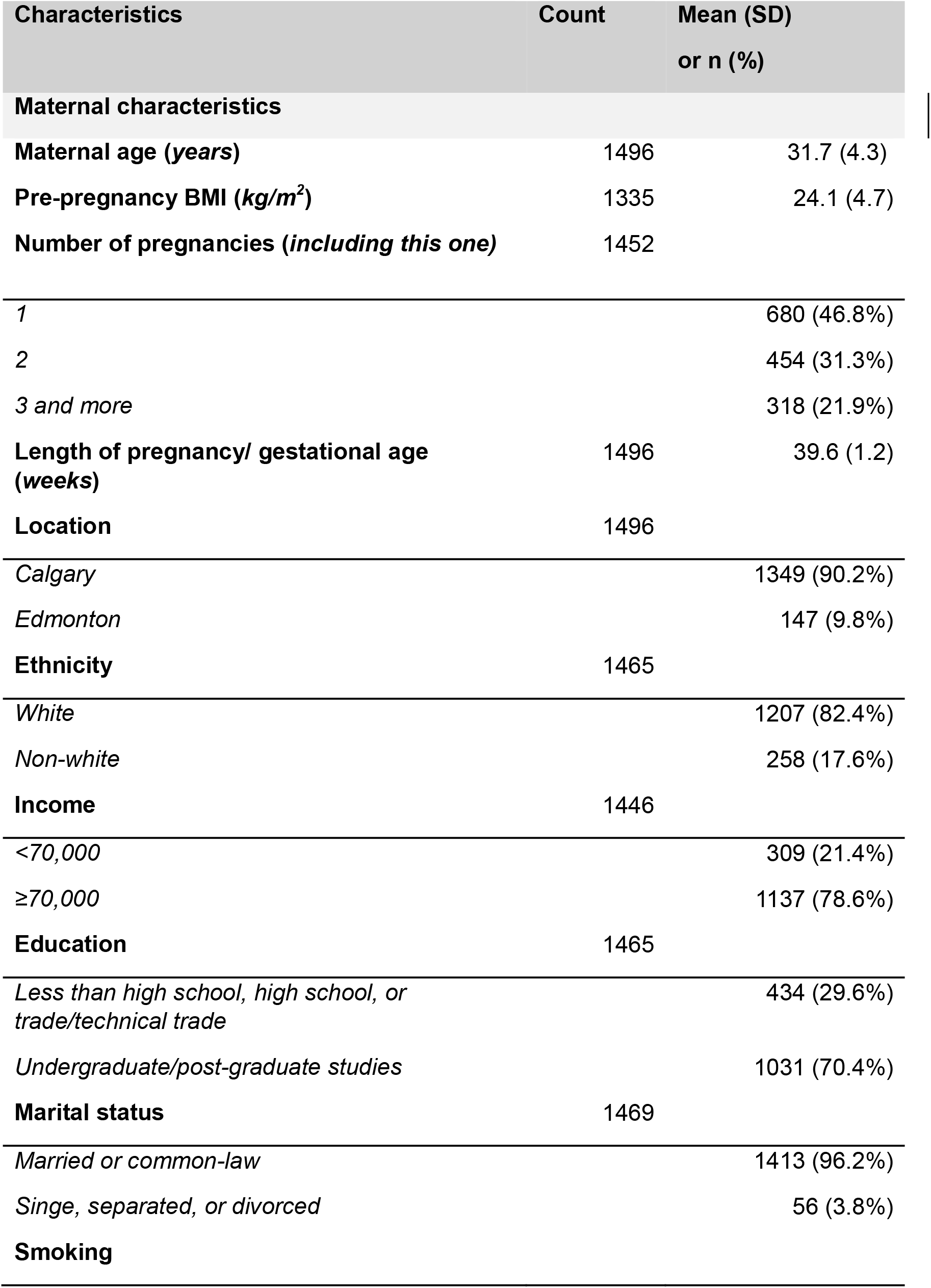
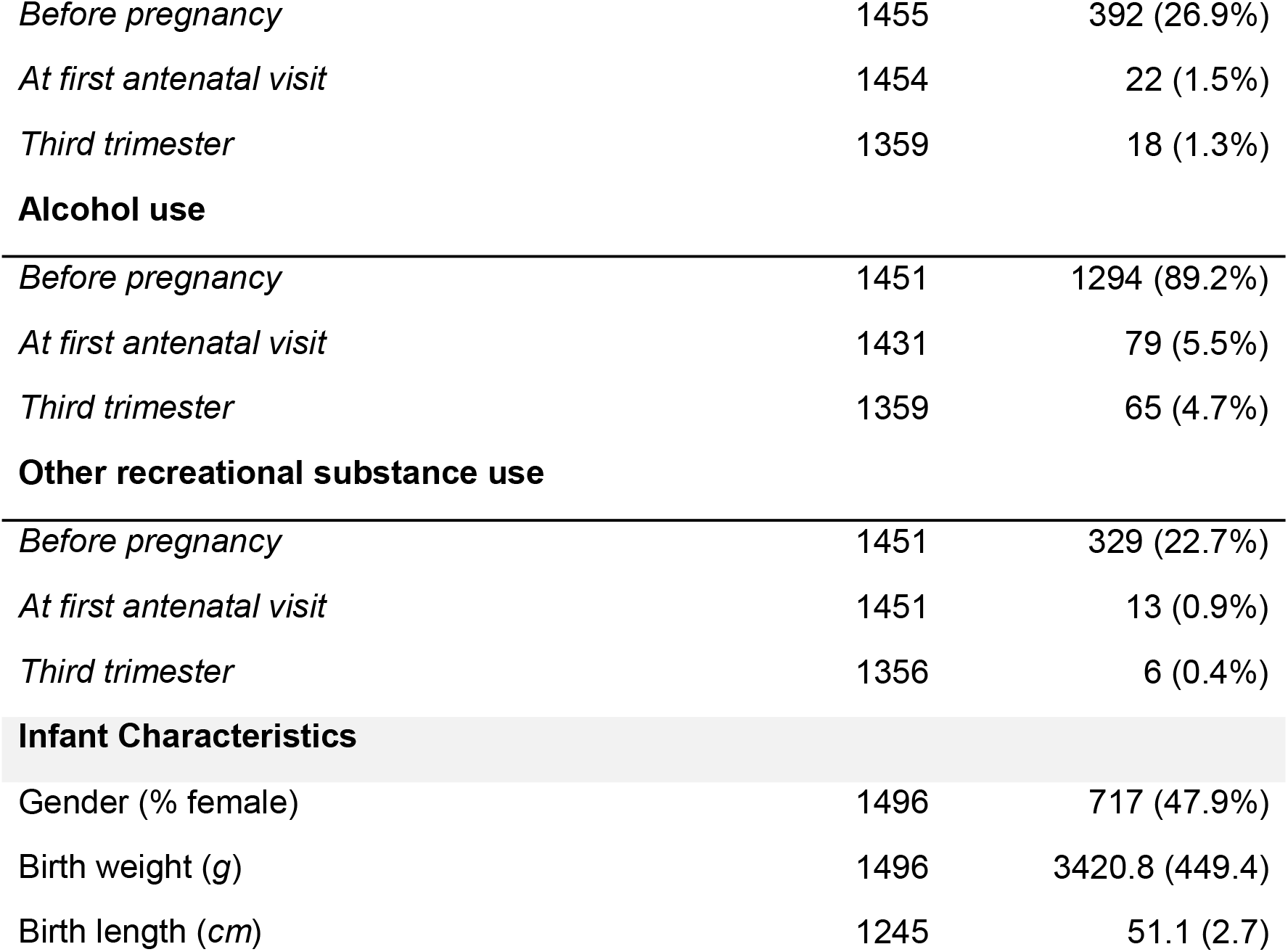
Descriptive statistics of 1496 maternal-infant pair participants in the APrON cohort.

| Characteristics | Count | Mean (SD)<br>or n (%) |
| --- | --- | --- |
| **Maternal characteristics** |  |  |
| **Maternal age (years)** | 1496 | 31.7 (4.3) |
| **Pre-pregnancy BMI ($kg/m^2$)** | 1335 | 24.1 (4.7) |
| **Number of pregnancies (*including this one*)** | 1452 |  |
| 1 |  | 680 (46.8%) |
| 2 |  | 454 (31.3%) |
| 3 and more |  | 318 (21.9%) |
| **Length of pregnancy/ gestational age (weeks)** | 1496 | 39.6 (1.2) |
| **Location** | 1496 |  |
| Calgary |  | 1349 (90.2%) |
| Edmonton |  | 147 (9.8%) |
| **Ethnicity** | 1465 |  |
| White |  | 1207 (82.4%) |
| Non-white |  | 258 (17.6%) |
| **Income** | 1446 |  |
| <70,000 |  | 309 (21.4%) |
| ≥70,000 |  | 1137 (78.6%) |
| **Education** | 1465 |  |
| Less than high school, high school, or trade/technical trade |  | 434 (29.6%) |
| Undergraduate/post-graduate studies |  | 1031 (70.4%) |
| **Marital status** | 1469 |  |
| Married or common-law |  | 1413 (96.2%) |
| Singe, separated, or divorced |  | 56 (3.8%) |
| **Smoking** |  |  |
| *Before pregnancy* | 1455 | 392 (26.9%) |
| *At first antenatal visit* | 1454 | 22 (1.5%) |
| *Third trimester* | 1359 | 18 (1.3%) |
| **Alcohol use** |  |  |
| *Before pregnancy* | 1451 | 1294 (89.2%) |
| *At first antenatal visit* | 1431 | 79 (5.5%) |
| *Third trimester* | 1359 | 65 (4.7%) |
| **Other recreational substance use** |  |  |
| *Before pregnancy* | 1451 | 329 (22.7%) |
| *At first antenatal visit* | 1451 | 13 (0.9%) |
| *Third trimester* | 1356 | 6 (0.4%) |
| **Infant Characteristics** |  |  |
| Gender (% female) | 1496 | 717 (47.9%) |
| Birth weight (g) | 1496 | 3420.8 (449.4) |
| Birth length (cm) | 1245 | 51.1 (2.7) |

**Table 2.** Descriptive statistics of maternal participant biomarkers across pregnancy.

|  | First trimester | Second trimester | Third trimester | Postpartum |
| --- | --- | --- | --- | --- |
| sFer |  |  |  |  |
| *n* | 312 | 1451 | 1325 | 1283 |
| *mean (sd)* | 41.8 (1.9) | 29.7 (2.1) | 13.5 (1.8) | 40.6 (2.1) |
| sFer adjusted for inflammation |  |  |  |  |
| *n* |  |  | 1251 |  |
| *mean (sd)* |  |  | 12.3 (1.8) |  |
| Hemoglobin |  |  |  |  |
| *n* | 319 | 1381 | 1319 | 1336 |
| *mean (sd)* | 128.5 (8.9) | 122.7 (8.5) | 122.3 (9.0) | 133.9 (9.0) |
| C-reactive protein |  |  |  |  |
| *n* | 35 | 130 | 1284 | 117 |
| *mean (sd)* | 6.7 (7.3) | 7.3 (5.8) | 3.6 (4.3) | 3.6 (4.1) |

**Figure 1.**
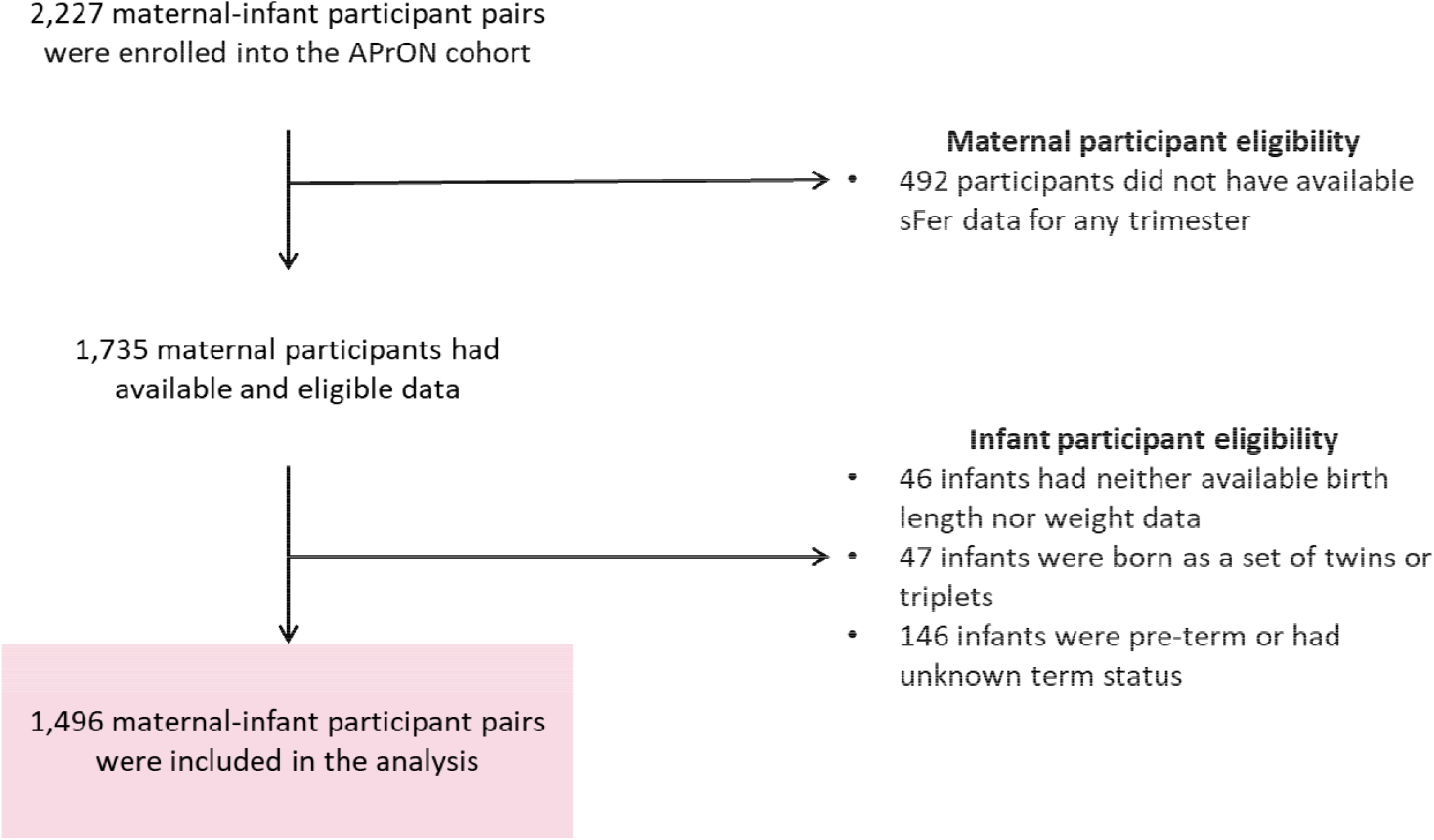
Flow chart of the total number of participants included in the analysis

**Figure 2.**
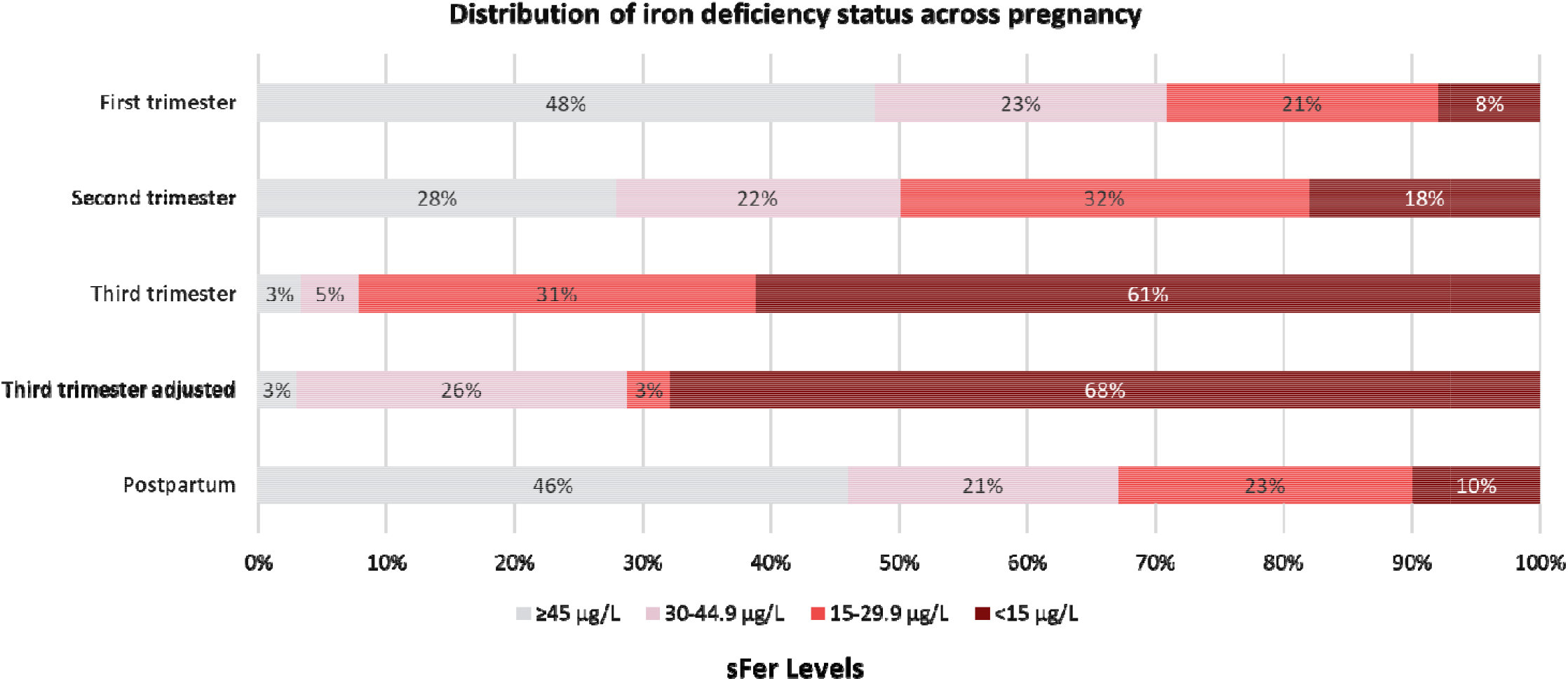
Distribution of iron deficiency status across pregnancy

### Bivariate analysis of maternal predictors on anthropometric birth outcomes

In unadjusted analysis, inflammation-adjusted serum ferritin in T3 was negatively associated with birth weight (p < 0.001) (Table 3). Serum ferritin in T2 and T3 were not associated with birth weight nor birth length. White ethnicity, higher income, higher number of pregnancies, higher BMI, longer gestational age, alcohol drinking before pregnancy were positively associated with higher birth weight (Table 3). Boys were on average 143g heavier than girls (p<0.001).

**Table 3.**
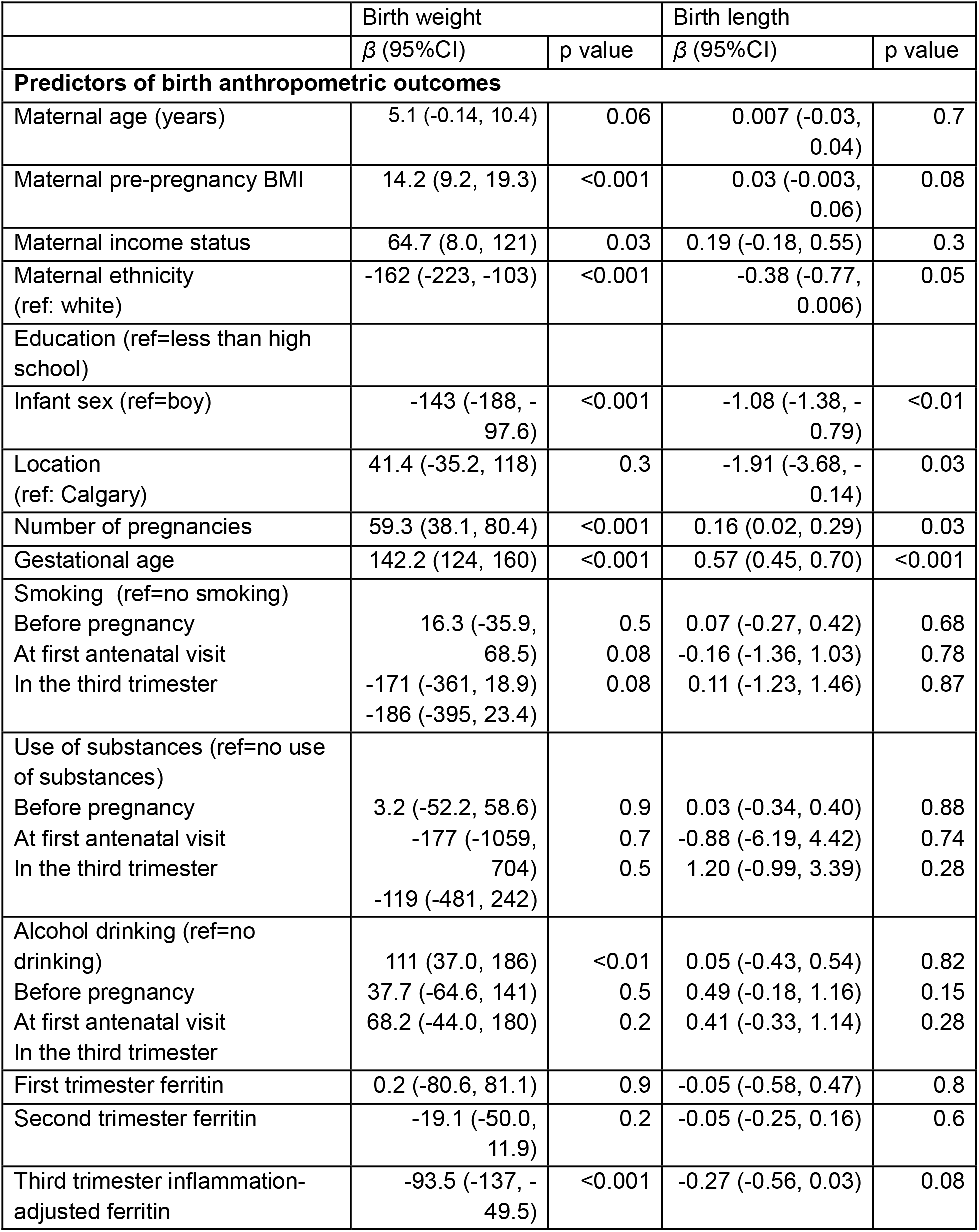
Bivariate analysis of maternal socio-demographic characteristics with infant anthropometric outcomes at birth.

### Multivariate analysis of maternal predictors on anthropometric birth outcomes

In a model adjusted for ethnicity, location, education, number of pregnancies, gestational age, maternal age, pre-pregnancy BMI, sex of the infant, use of substances before and after pregnancy, lower inflammation-adjusted serum ferritin remained significantly associated with higher birth weight (Table 4). Specifically, each 1 unit increase in log ferritin was associated with a 93.3 g lower birth weight (β = – 93.3 g; 95% CI: –136 to –51, p < 0.001). This corresponds to approximately a 93.3 g higher birth weight for each 2.7-fold decrease in ferritin concentration.

**Table 4.**
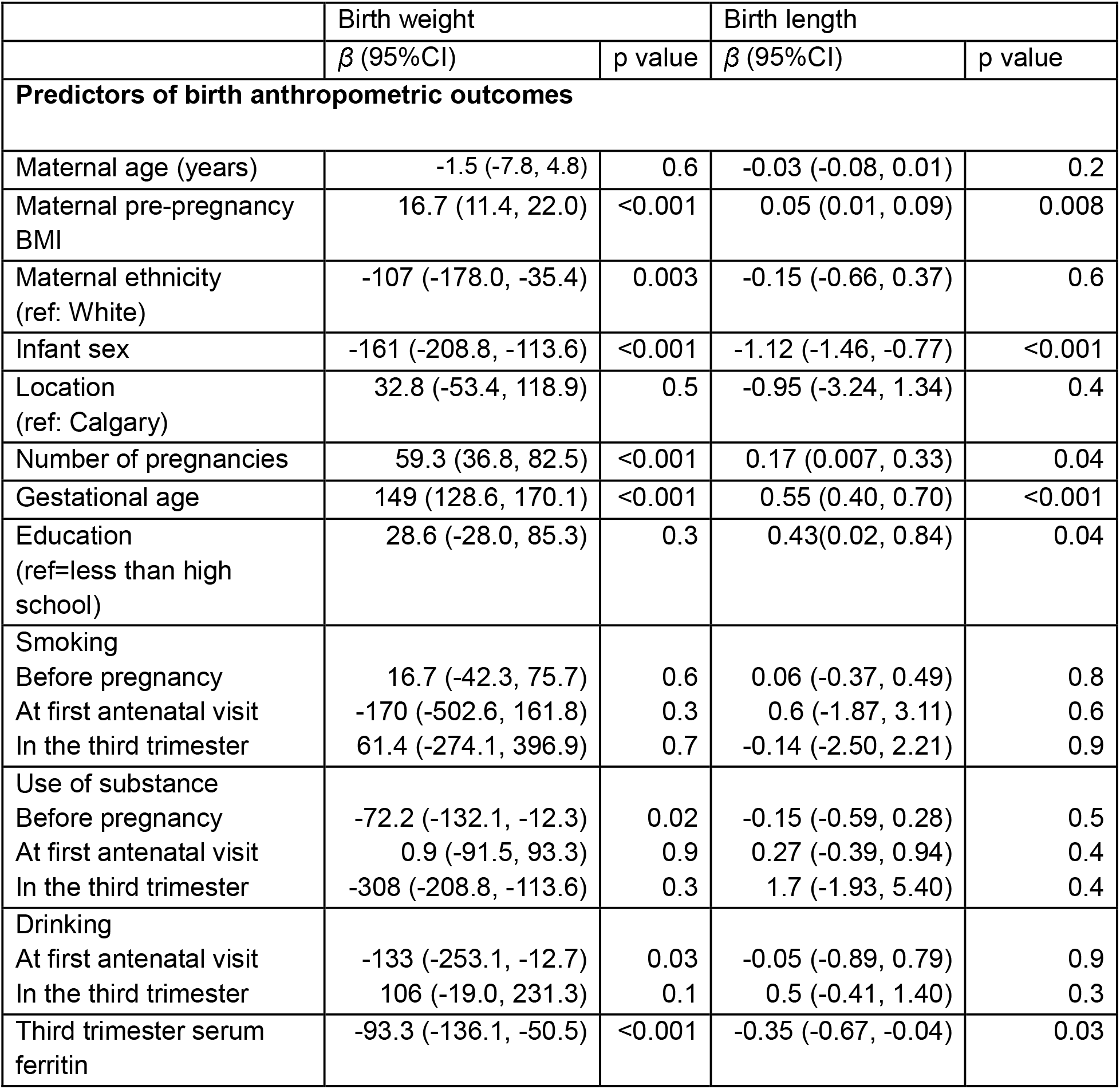
Multivariate analysis of maternal third trimester adjusted serum ferritin with infant anthropometric outcomes at birth.

Lower inflammation-adjusted T3 ferritin was also associated with greater birth length. A 1-unit increase in log ferritin was associated with a 0.35 cm shorter birth length (β = –0.35 cm; 95% CI: –0.66 to –0.04; p = 0.03), corresponding to a 0.35 cm longer birth length for each 2.7-fold decrease in ferritin concentration.

In the fully adjusted model, use of substance before pregnancy and alcohol drinking at the first antenatal visit were associated with lower birth weight.

All models satisfied assumptions of linearity, homoscedasticity, and multicollinearity.

### Longitudinal analysis

Among women with paired T2 and T3 ferritin values, we examined Δlog ferritin (T2–T3) to quantify late pregnancy iron mobilisation. Mean birthweight differed by the magnitude of change in ferritin between T2 and T3 (p = 0.002). Compared with women who experienced a large reduction in ferritin concentrations, birthweight was on average 155 g lower in those with no or positive change (β = −155.0 g, 95%CI: −248, −62, p = 0.001), and 103 g lower in those with a modest reduction in iron stores (β = −102.6 g, 95%CI: −171, −34, p = 0.003) (Figure 3).

**Figure 3.**
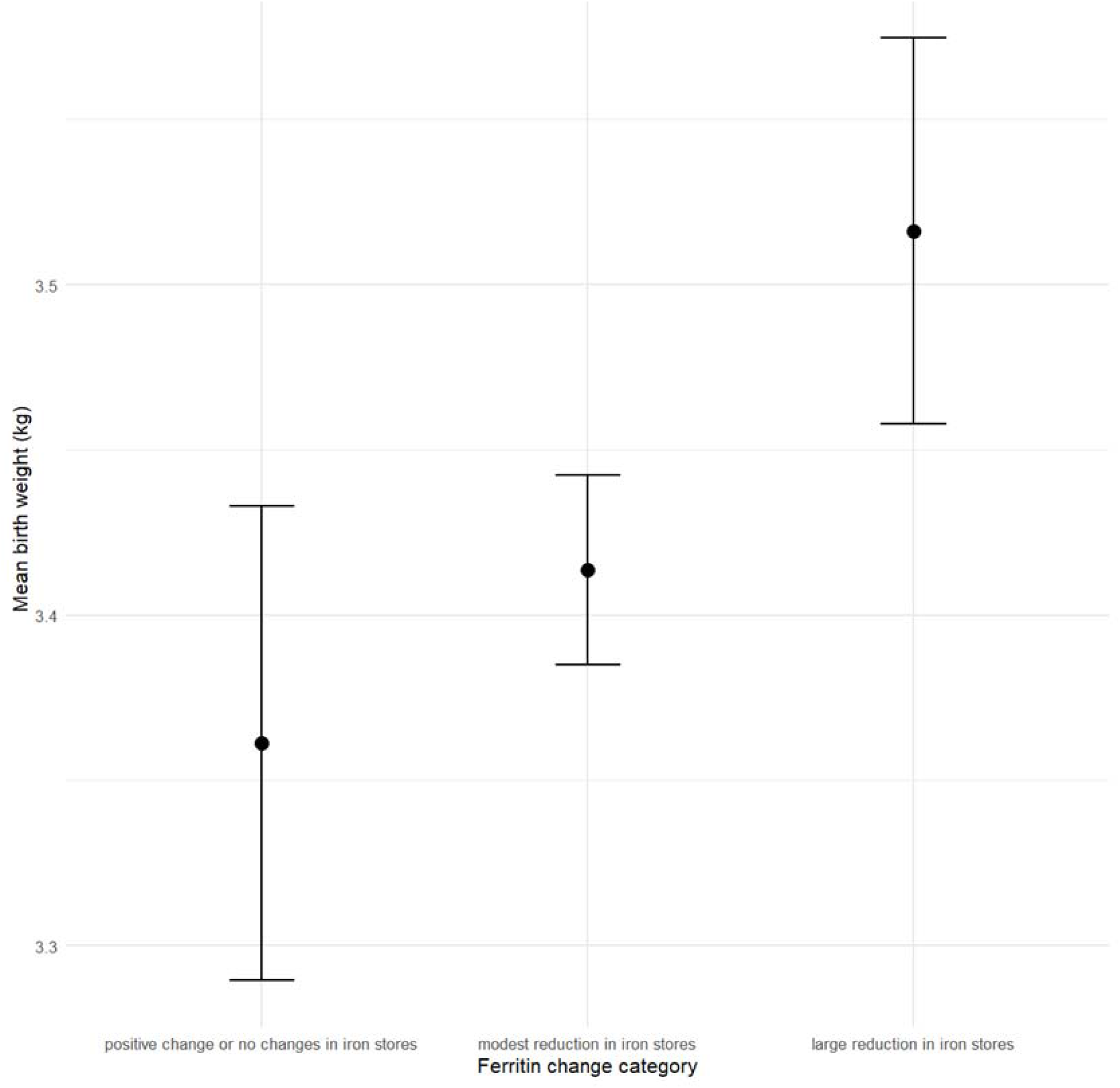
Mean infant birth weight (in kg) by level of changes in maternal serum ferritin between the second and third trimesters

The association remains significant after adjustment for maternal age, education, ethnicity, location, gestational age and inflammation in the third trimester, but not after adjusting for maternal BMI, suggesting partial confounding by maternal body size.

### Post-partum analysis

Postpartum ferritin showed no association with birthweight or birth length in linear models. However, spline analyses indicated a non-linear threshold pattern (P < 0.001), with higher birthweight and length observed at the lowest ferritin concentrations and little variation across moderate or higher levels.

In adjusted logistic regression models, each 1 kg increase in birthweight was associated with 83% higher odds of postpartum ID (OR 1.83; 95% CI 1.12–2.99; p=0.02). Each 1 cm increase in birth length was associated with 12% higher odds (OR 1.12; 95% CI 1.03–1.23; p=0.01).

## Discussion

In this longitudinal cohort of healthy, predominantly high-income women, we evaluated the timing and dynamics of iron status during pregnancy and postpartum, and their associations with infant anthropometric outcomes. We found that lower maternal ferritin concentrations in late pregnancy were associated with higher birthweight. We also observed that women who experienced the greatest declines in ferritin between mid and late pregnancy delivered the largest infants. Finally, at three months postpartum, women with the lowest ferritin concentrations had given birth to the heaviest and longest infants. These findings reveal a pattern in which maternal iron depletion may reflect increased foetal iron transfer rather than impaired growth in this low-risk population, and highlight the substantial iron demands of late gestation, particularly when carrying a large foetus.

Our findings contrast with much of the global literature, which typically reports that maternal ID and anaemia are associated with lower birthweight, preterm birth, small for gestational age infants, and reduced neonatal size (21). However, those studies often rely on haemoglobin and are conducted in settings where ID is severe, anaemia is widespread, and coexisting nutritional and infectious burdens influence haemoglobin, ferritin and foetal growth (22,23). In contrast, our cohort had low anaemia prevalence and low inflammation, and ferritin values therefore are more likely to reliably reflect iron stores.

Emerging evidence supports the pattern observed in our study. A Danish study similarly found that lower, non-adjusted ferritin in pregnancy was associated with higher birthweight z-scores (28). These results align with experimental isotope studies demonstrating that when maternal iron stores decline, the placenta increases its efficiency of iron transfer to the foetus (24,25). This mechanism ensures foetal iron accretion even at the expense of maternal reserves and is most pronounced in late gestation, when foetal iron uptake accelerates. This is consistent with documented iron physiology during pregnancy and the established association between infant iron levels in relation to their bodyweight. Dallman et al. have estimated that at 20 and 40 weeks gestation, the foetus contains about 58 and 94 µg of iron respectively per gram of lean tissue. It is also established that iron endowment occurs during the third trimester (26), a critical time when the foetus accumulates about 2 mg of iron daily (15).

We observed that the largest drops in ferritin between the second and third trimester occurred among women who delivered the largest infants, supporting the mechanism that iron endowment is proportional to their body weight, at the expense of maternal iron stores. Importantly, haemoglobin levels remained normal, indicating that erythropoiesis was preserved even as iron stores were being mobilized. This is supported by other evidence showing that reduction of serum ferritin may signal iron depletion without iron erythropoiesis being thwarted, as iron is prioritized for haematopoiesis (27). Iron supply to the foetus is favoured by changes in the structural and compositional characteristics of transferrin which facilitate preferential delivery of iron to placental rather than systemic transferrin receptors (28).

Our postpartum findings further reinforce this interpretation. At three months postpartum, maternal ferritin concentrations had returned to values similar to the first trimester, and haemoglobin levels were normal. Nevertheless, women who had delivered larger infants remained iron-depleted, as reflected in lower postpartum ferritin. This pattern is best explained by persistent depletion of maternal iron reserves following increased foetal iron transfer during late gestation, particularly in heavier infants.

A longitudinal study in healthy pregnant women found that dietary absorption of iron increased from 7% at 12 weeks to 36% and 66% at 24 and 36 weeks gestation respectively; at 16–24 weeks postpartum, absorption decreased to 11% (29). This shows that, while maternal mechanism are in place to increase iron absorption and transfer to the foetus during pregnancy, maternal iron absorption decreases postpartum, which may limit rapid repletion of maternal iron stores.

This interpretation has important implications for clinical care. First, women who deliver larger infants may be at greater risk of postpartum iron deficiency—even if not anaemic—and may benefit from targeted iron supplementation after delivery. Second, routine haemoglobin screening during pregnancy is insufficient, as haemoglobin remained normal even in women with substantial ferritin decline. Monitoring ferritin across pregnancy offers valuable information about maternal iron dynamics and foetal iron accretion, particularly during the third trimester, when iron endowment to the foetus peaks.

Finally, our findings underscore the need for improved iron screening and management guidelines during pregnancy and postpartum. Despite low anaemia prevalence, ID was highly prevalent and clinically relevant in this cohort. It is worth noting that, unlike the UK and the EU where reference intakes for iron during pregnancy are the same as those of non-pregnant women, reference intakes for iron during pregnancy are 1.5 times more than that for non-pregnant women in Canada (15).

Addressing maternal iron depletion requires both antenatal monitoring and postpartum follow-up, particularly for women with rapidly falling ferritin or those who deliver large infants.

### Strengths and Limitations

This study has several important strengths. First, the longitudinal design allowed assessment of ferritin concentrations across multiple time points in pregnancy and postpartum, enabling evaluation of iron trajectories rather than single measurements. Second, anaemia prevalence and inflammation were low in this cohort, improving the interpretability of ferritin as a marker of iron stores. Furthermore, our sample was sufficiently large, with a data being drawn from a sample of 1496 maternal-infant participants,

Missing data present an important limitation. Birth length was missing for approximately 250 infants, reducing statistical power for models using this outcome. In addition, inflammation adjusted ferritin could only be computed for women with available CRP measurements, resulting in the exclusion of 74 participants from adjusted ferritin models. In addition, maternal BMI was missing for 161 women. This reduction in sample size likely contributed to the loss of statistical significance for the association between the change in ferritin concentration in late pregnancy and birth weight, despite a similar direction and magnitude of effect to the model that was not adjusted for maternal BMI.

The cohort was predominantly white, well-educated, and high-income, which limits generalisability to more diverse or higher risk populations. In our study, participants who were a Person of Colour or were of low income delivered slightly smaller infants, consistent with evidence that social and structural disadvantage is associated with lower birth weight, shorter gestational age, and higher risks of preterm birth and neonatal mortality (30,31). As such, we may may not have sufficiently captured the populations who experience these suboptimal health outcomes at higher levels and require services the most. However, despite biases within our cohort, this analysis finds that even among high-income and well-educated mothers ID is found at high levels, indicating quality care and support must be given to all pregnant women regardless of sociodemographic and economic background.

We used ferritin to identify iron status without adjusting for inflammation at all time points, which may underestimate the prevalence of ID. However, a threshold of >19 mg/L during pregnancy is indicative of elevated inflammation, indicating that the influence of inflammation is likely low in our cohort (32). Nevertheless, this study provides a novel exploration of the relationship between ID across pregnancy and postpartum, evaluating sFer in a longitudinal manner. This study generates new insights into understanding the complexities of ID during pregnancy in a high-income setting.

## Conclusion

In this longitudinal cohort of healthy pregnant women, lower ferritin concentrations and greater declines in ferritin during late gestation were associated with higher birthweight. These findings are consistent with preferential foetal iron accretion during the third trimester. Further studies in more diverse settings are needed to determine whether similar patterns are observed in populations with higher inflammation burdens or greater anaemia prevalence.

## Data Availability

Information about the APrON study is available at www.apronstudy.ca. APrON data are available through Secondary Evidence to Generate Evidence (https://policywise.com/sage/). For more information, contact Principal Investigator, Dr. Nicole Letourneau at. Collaboration or data access inquiries will be considered by the APrON Study team.

## Author contribution

PK: conceptualisation, data curation, formal analysis, writing - original draft. FS: conceptualisation, data curation, formal analysis, methodology, supervision, writing - original draft, writing - review and editing. RB: conceptualisation, funding acquisition, project administration, supervision, writing - review and editing. SB: conceptualisation, project administration, supervision, writing - review and editing.

